# Technology-delivered undergraduate medical education involving patients and carers: A rapid systematic review

**DOI:** 10.1101/2021.05.07.21256812

**Authors:** Sadie Lawes-Wickwar, Eitan Lovat, Adedoyin Alao, Julia Hamer-Hunt, Nesrin Yurtoglu, Cherise Jensen, Nicola Clarke, Nia Roberts, Sophie Park

## Abstract

**Background:** Involving patients and carers in medical education centralises their voice in healthcare and supports students to develop key professional and person-centred skills. Medical schools are increasingly using technology to deliver educational activities. No review currently exists to establish the variety of technologies and their uses in undergraduate medical education when patients and/or carers are involved.

**Methods:** Ovid MEDLINE, Ovid EMBASE and medRxiv were searched in October 2020 and reference lists of key articles were hand searched. Eligible studies reported technology-assisted education, in any setting, involving authentic patients and/or carers. Studies in foreign languages, or describing actors or non-authentic patients were excluded. Study quality was assessed using the Mixed Methods Appraisal Tool (MMAT). Levels of patient involvement were assessed using Towle et al’s (2010) taxonomy.

**Results:** Twenty studies were included. The majority involved patients and/or carers via pre-recorded videos or online scenarios, with no student-interaction. Four studies evaluated remote consultations using telehealth technology, involving real-time interactions with authentic patients. Technology-supported teaching sessions involving patients and/or carers were found to be acceptable to students, educationally valuable (to students and educators), and enhanced student engagement, patient-centred attitudes, knowledge of specific patient groups, and communication and clinical skills. Two studies describing real-time remote interactions with authentic patients indicated potential barriers for students (reduced ability to build relationship with patients and examine them), educators (reduced ability to build rapport with students) and patients (issues with using or accessing telehealth).

**Conclusions:** No studies directly measured the perspective of patients or carers involved in technology-delivered medical education. Future research should establish barriers and facilitators to patients and carers taking up a role in medical students’ education when technology is used, and evaluate PPI activities at Levels 3 and above as described by Towle et al’s taxonomy.

## 1. INTRODUCTION

Patient-centred healthcare is now at the forefront of the NHS, and personalised care is central to the NHS Long Term Plan [1]. To achieve this, it is important that medical education centralises the experiences and perspectives of patients and their carers. Patient and public involvement (PPI) in undergraduate medical education is recommended by the General Medical Council (GMC), in roles including teaching, assessment, feedback, and curriculum development [2]. PPI in medical teaching can be a driver towards more person-centred care as students move into professional practice [3]. Other widely reported benefits include: improved professional attitudes and empathic communication skills among students, improved clinical performance as both students and medical professionals, as well as professional, personal, and emotional benefits for involved patients and carers [3–4].

PPI is an increasingly common feature of healthcare education, however what is considered “involvement” varies widely. A recent taxonomy of involvement in health education defines active PPI as a spectrum, from featuring in case studies (“Level 1”) to involvement at an institutional level, and involved in decision-making (“Level 6”) [5]. Historically, patients and the public have held relatively passive roles in the education of medical students, but examples of good practice have increased over recent years [6]. Other than examples relating to electronic case studies, or pre-recorded videos of patients or carers viewed by healthcare students [5–7], the variety of uses of technology in the delivery of medical education has not been widely considered by recent PPI frameworks.

The ongoing COVID-19 pandemic has presented challenges for medical teaching; newly implemented rules regarding social distancing has led to a reduction of in-person teaching and an increase in remote learning [8]. Despite this shift, the involvement of patients and carers should continue to support students’ professional and communication skills, help them respond to the evolving needs of patient groups [9], and develop skills in consulting remotely; a key requirement for the future healthcare workforce [10, 11]. However, barriers might arise from this new way of working, for example where patients or carers do not have access to a reliable internet connection [12]. “Digital exclusion” describes the consequences of barriers to accessing remotely-delivered healthcare. For example, mental health service users report being unable to access necessary technology, and finding connecting remotely more difficult due to their symptoms, in recent research [13].

As far as we are aware, no existing reviews have reported the variety of technologies and their uses to deliver educational activities involving patients and carers in undergraduate medical education. Two recent systematic reviews have described active PPI in medical education broadly [3, 14], however these reviews were not focused on the use of technology. Another recent scoping review found telephone and internet technologies were the most commonly utilised forms of technology when involving patients in rural healthcare education settings [15], where patients were consulted about new curricula, evaluated programmes and their views about the use of healthcare technology were measured [15]. However, this scoping review was limited to rural healthcare and was not specific to undergraduate medical education. The aims of the present review were to 1) identify the use and evaluation of technology in undergraduate medical teaching when patients and/or carers have been involved, encompassing all educational settings and geographical locations; 2) identify the levels of patient and/or carer involvement when technology is used, and 3) identify reported barriers and facilitators to PPI when technology has been used.

## 2. METHODS

Rapid systematic review methods were employed. Rapid reviews follow standard systematic review procedures, whilst providing timely evidence and maintaining rigour [16]. Rapid methods were chosen to provide teaching teams with timely evidence for the uses of technology to support continued PPI in undergraduate medical education after the rapid shift to remote working during the COVID-19 pandemic.

### 2.1 Protocol

The protocol has been registered on PROSPERO Ref. CRD42021243279. 2.2 Search strategy and selection criteria Searches for published and unpublished studies were performed from database inception to 27th October 2020 using MEDLINE (OvidSP), EMBASE (OvidSP) and medRXiv Preprints (https://www.medrxiv.org/). The search strategy is available as a supplementary file (Supplementary File 1). Searches were not limited by language or publication date. Retrieved references were initially de-duplicated in Endnote before being exported into Rayyan [17] and titles and abstracts were screened. Ten percent of titles and abstracts were screened independently by two authors and any disagreements were discussed until consensus was reached.

Primary studies evaluating undergraduate medical education activities, using any type of technology (e.g. video, telephone, video-conferencing software, website) to involve patients and/or carers at any level [5], and employing any study design, were eligible. Eligible studies also reported student-, educator- and/or patient-related outcome data. Studies describing the use of actors (without experience of the medical problem they were presenting with) or other persons not presenting as authentic patients or carers were excluded. Non-English language articles were excluded due to the rapid nature of this review and a lack of resources to translate studies. Attempts were made to retrieve articles from authors’ institutions but if unsuccessful the article was excluded, due to time and funding restrictions. Reviews were excluded, but reference lists were hand searched for additional studies.

### 2.3 Data extraction and analysis

A data extraction form was developed by the authors based on the Sample, Phenomenon of Interest, Design, Evaluation, Research type (SPIDER) criteria, developed for reviewing qualitative and mixed methods studies [18]. A narrative synthesis was performed. A taxonomy of active PPI in healthcare education [5] was used to categorise the level of patient and/or carer involvement in the educational activity described by study authors. Categories range from patients being involved in developing a case study/ scenario - but had no overall influence on the theme of the content, nor on curriculum development (Level 1) - to patients being involved at the institutional level (Level 6) [5].

### 2.4 Quality assessment

The Mixed Methods Appraisal Tool (MMAT [19]) was used to assess study quality. The MMAT has been used for most common study methodologies and in a variety of contexts including health sciences, education, information sciences and psychology [19]. Two authors were independently involved in the appraisal process. MMAT scores were categorised as low, moderate or high-quality using criteria employed for two recent rapid systematic reviews of public health interventions [20, 21]; a score of 0-1 was categorised as low quality, 2-3 moderate quality, and 4-5 high quality.

## 3. PATIENT AND PUBLIC INVOLVEMENT (PPI) IN THE RESEARCH TEAM

The review team included two public contributors (JHH, NY), who joined the team at the stage of planning (after the research question had already been defined) and supported the review processes including literature screening, data extractions, and preparing the manuscript for publication. PPI contributors informed decisions about our inclusion criteria, ensuring the review considered involved carers (and not just patients).

## 4. RESULTS

### 4.1 Study selection

The full texts of 216 potentially relevant articles were screened for eligibility. A total of 20 studies were identified as eligible and included in the review (Figure 1).

**Figure 1.**
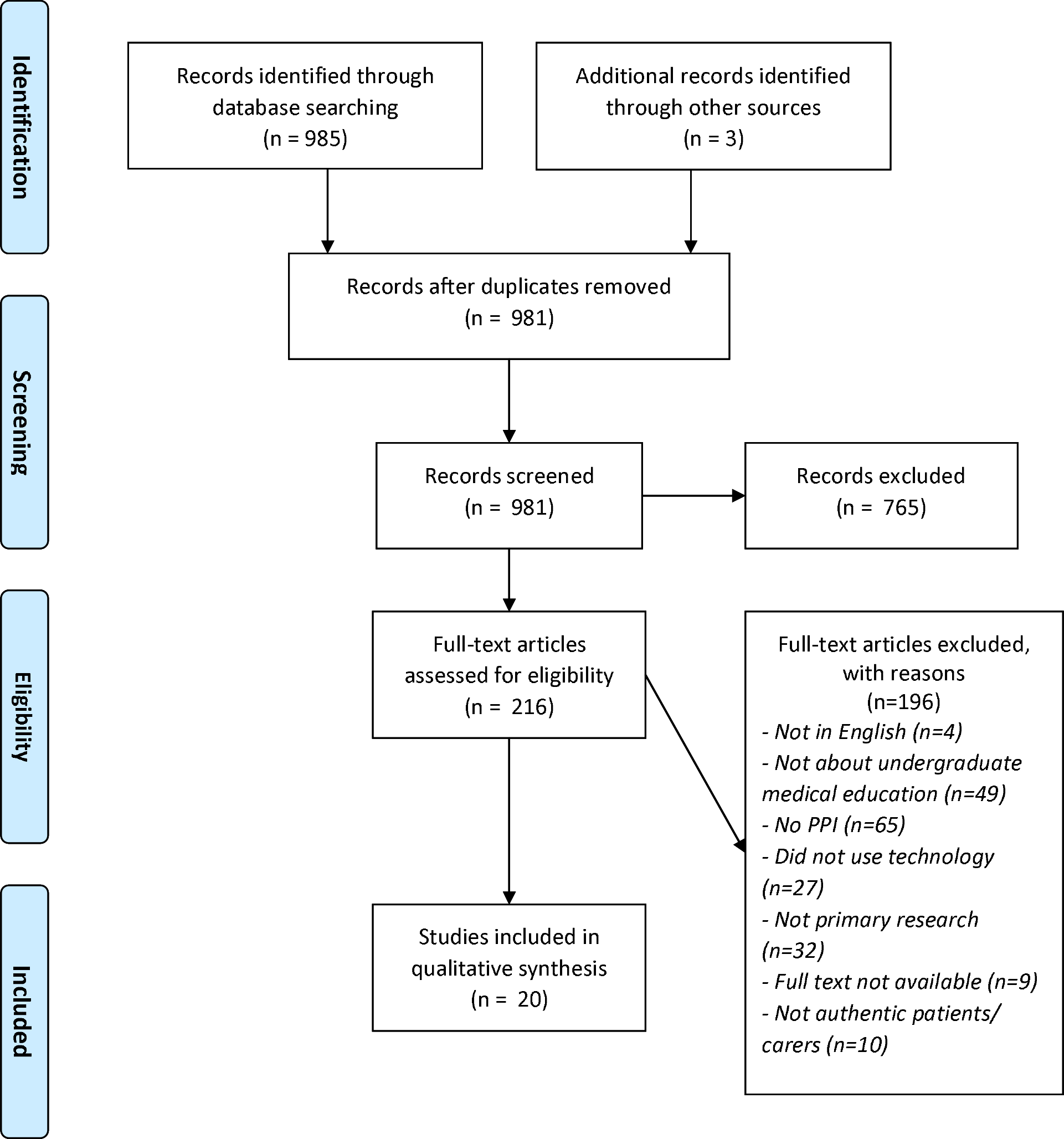
PRISMA Flow Chart

### 4.2 Study characteristics

Characteristics of the articles included in the review and the types of technology-supported educational interventions involving patients and carers are presented in Table 1.

**Table 1.**
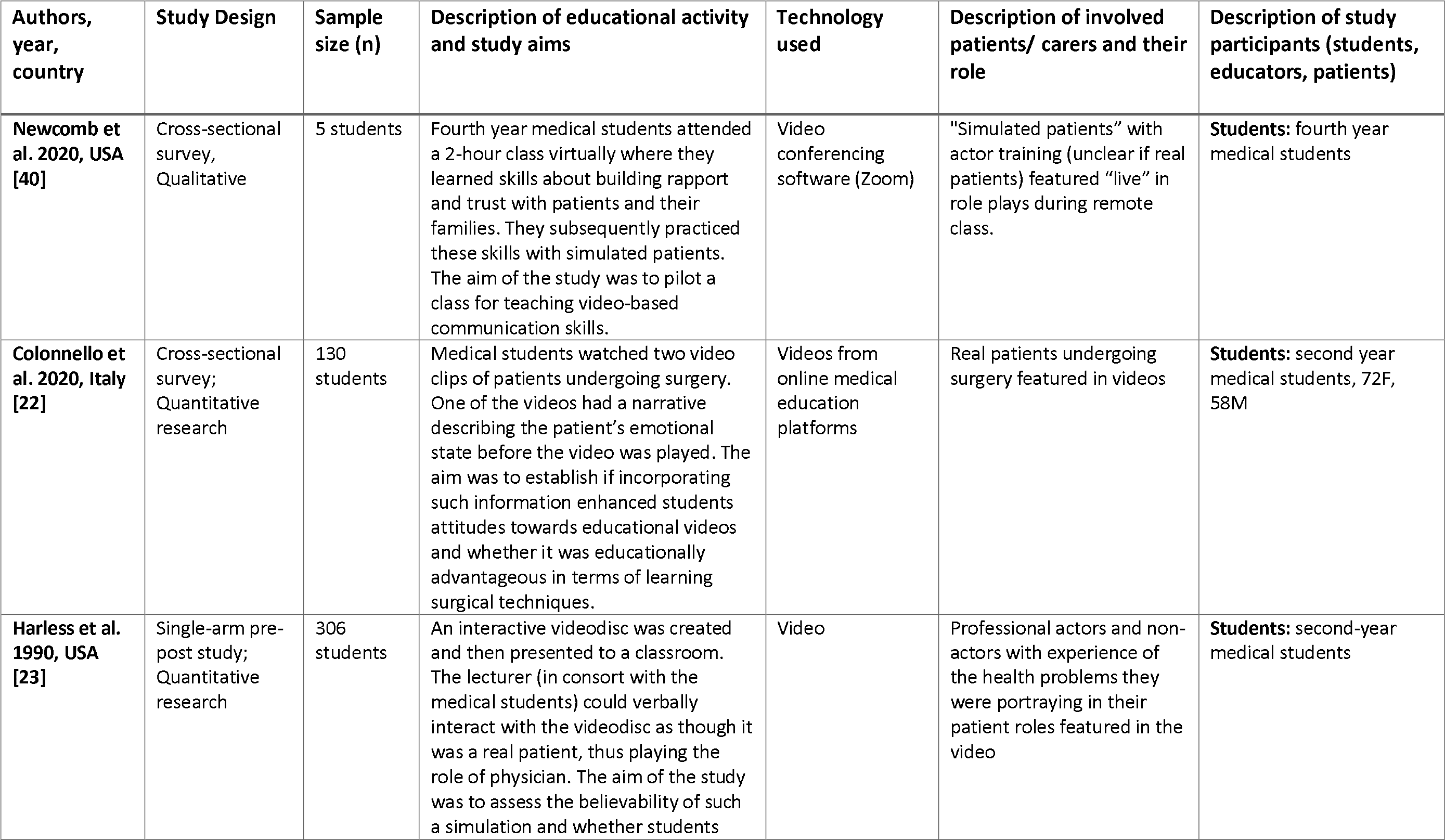
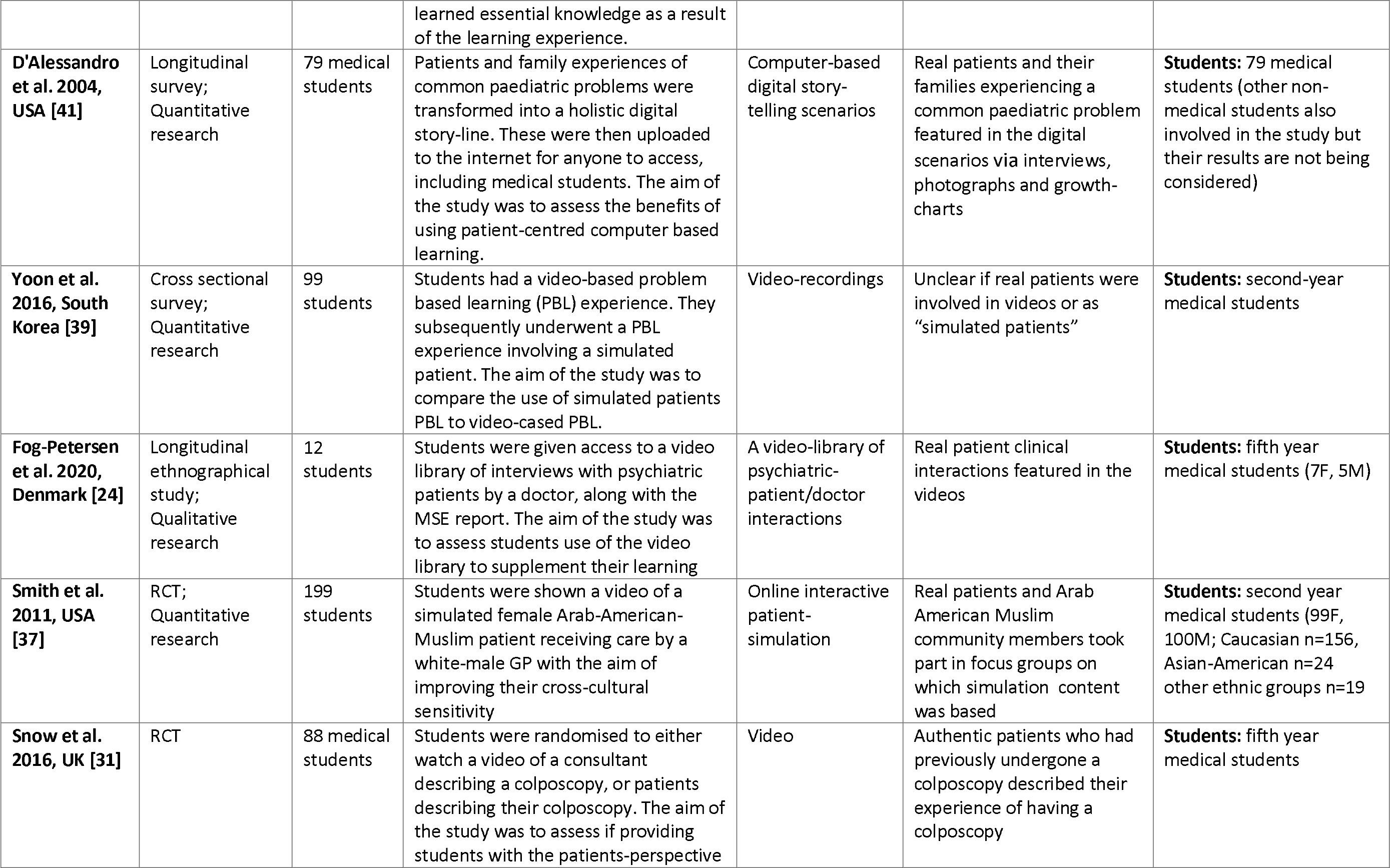
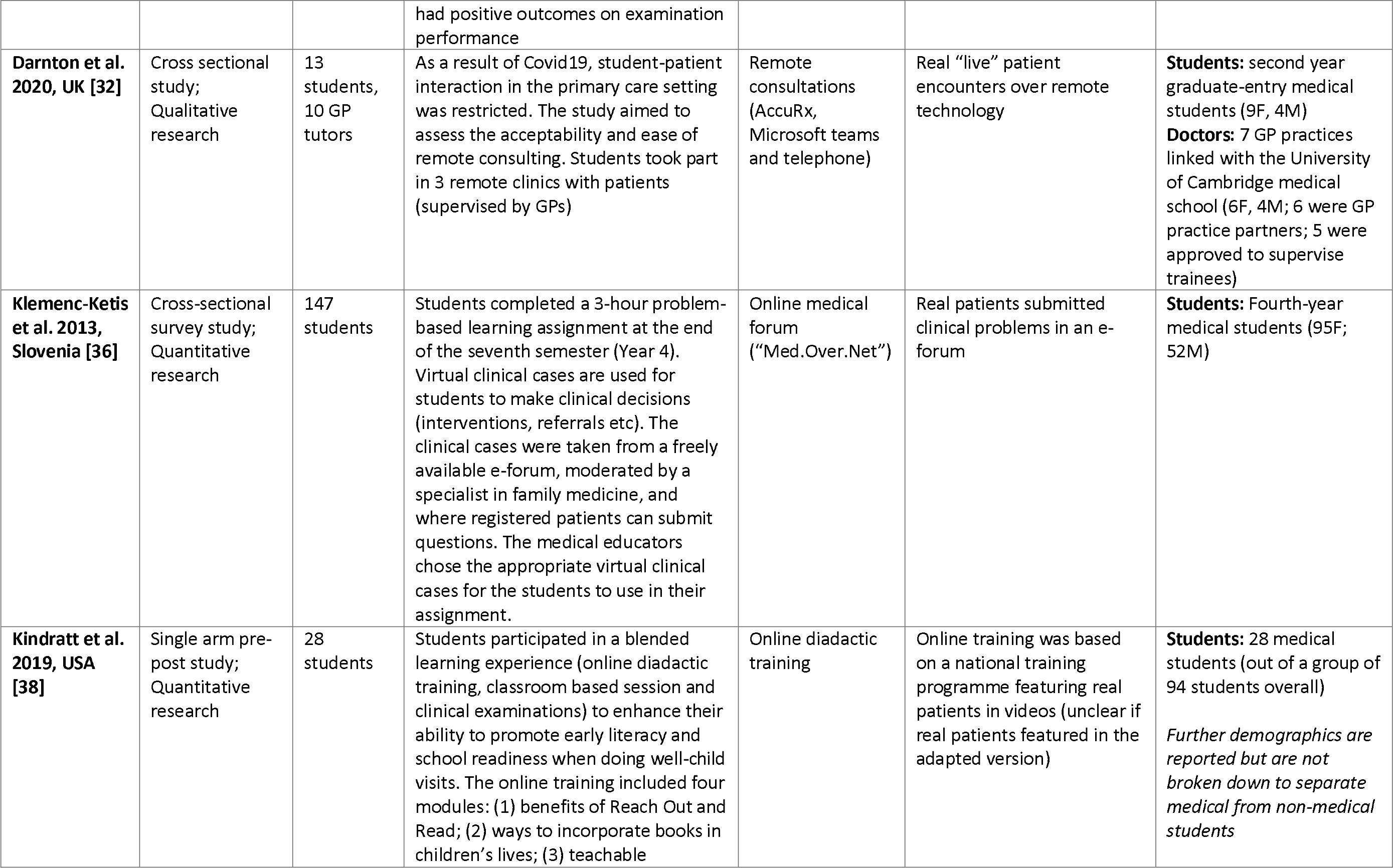
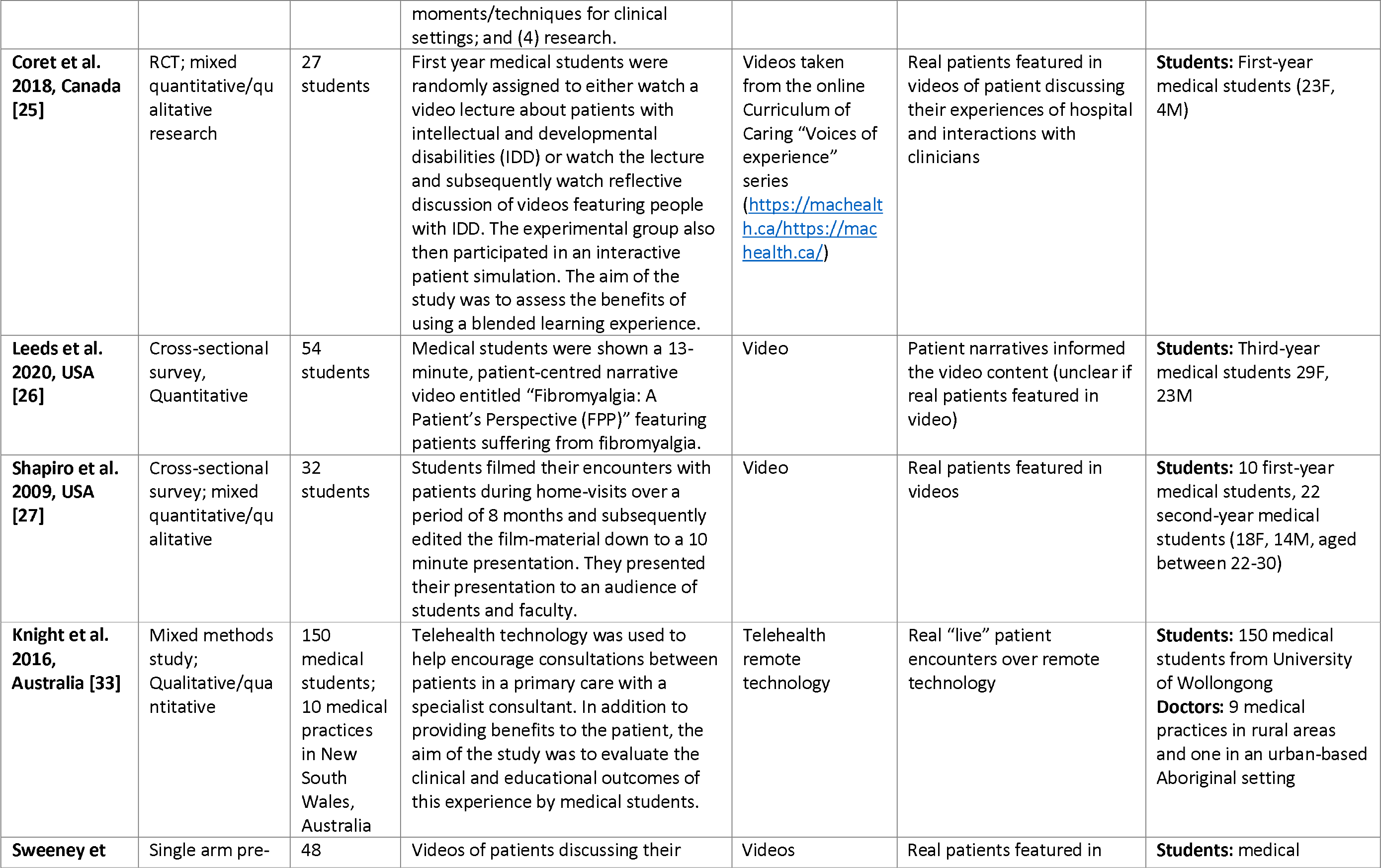
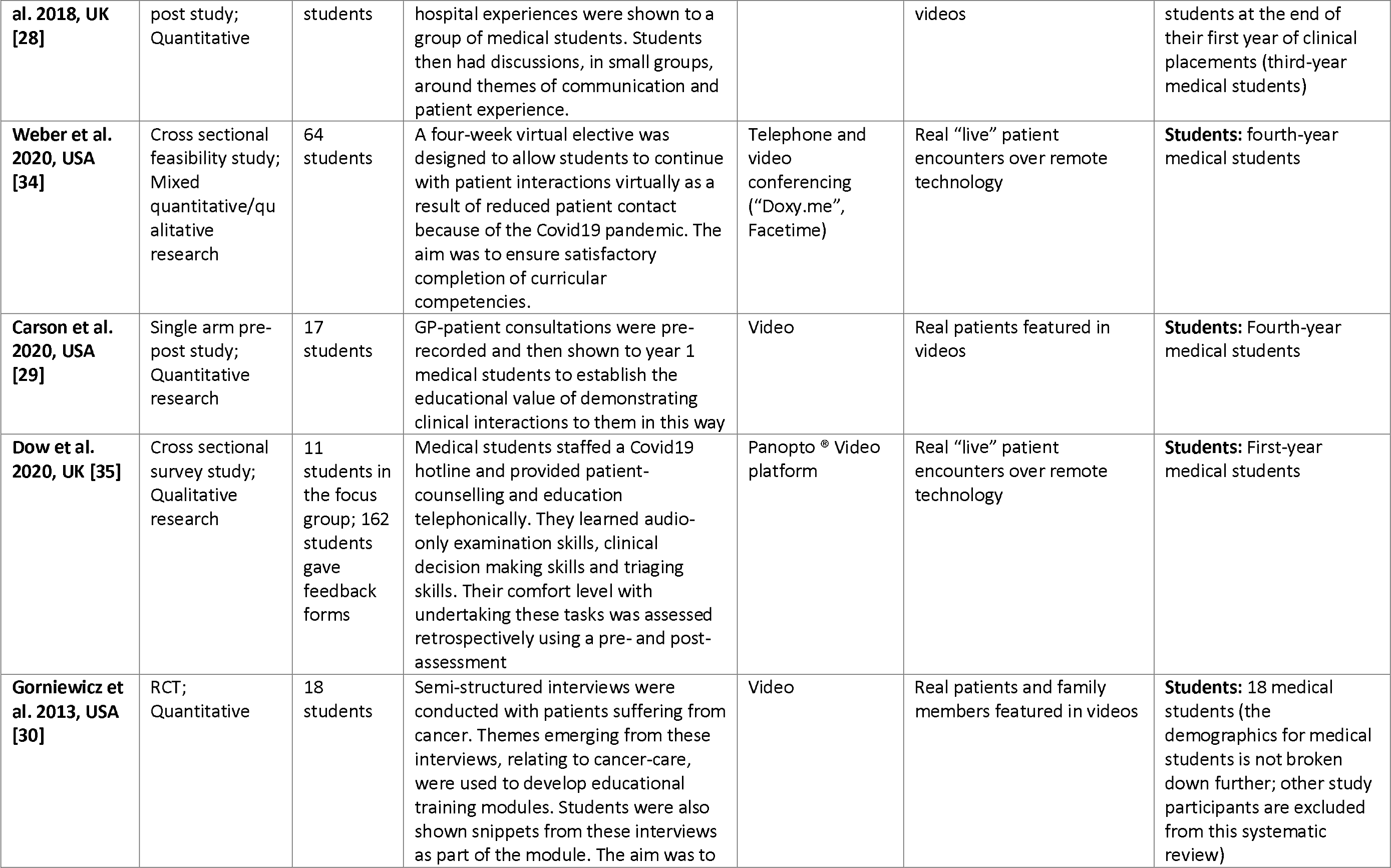
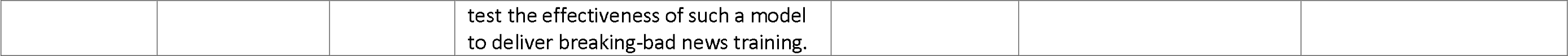
Characteristics of included studies

#### 4.2.1 Types of technology used

Ten out of the 20 included studies used pre-recorded videos to provide the patient’s perspective on their illness or demonstrate doctor-patient consultations [22–31]. Four studies involved remote consultation technology for student involvement in clinical consultations using telehealth platforms, video-conferencing software, and telephone [32–35]. Three studies used online technology, including an online medical e-forum where registered patients submitted questions [36], an online interactive patient simulation tool [37], and online didactic training [38]. One study compared the educational benefits of a video-lecture (the technology arm) to the use of “simulated” patients, where it is unclear what type of technology was used to simulate patients [39]. One study involved a class delivered via video-conferencing software where “simulated” patients were invited to feature in role plays with students [40]. One study used computer-based digital story boards to present common paediatric problems [41].

#### 4.2.2 Description of patients and carers involved in medical education

Two of the final 20 studies described using “simulated” or “virtual” patients, [39–40] but did not clarify what these terms meant. Two studies described patient-focused videos [26, 38], however a lack of description inhibited establishing whether patients were authentic. The remaining sixteen studies involved authentic patients [22-25, 27-37, 41]. One study, by Harless et al [23] employed professional actors and non-actors who all had experience with the health problems they were portraying in patient roles. Four studies evaluating remote consultations using telehealth technology involved “live” interactions with authentic patients [32–35]. The remaining studies involved patients in the development of materials that featured later in medical teaching (via technology). Two studies included the perspective of family members, one involved authentic family members [30] and one did not provide enough detail to determine if caregivers were authentic [38].

#### 4.2.3 Levels of patient and public involvement (PPI) in medical education

In the majority of cases, students viewed a pre-recorded video or completed online material that involved no real-time encounters with patients. These fourteen studies described patient and carer involvement categorised at Level 1 of Towle and colleagues’ [5] taxonomy [22-31, 36-38, 41]. Six studies involved patients in real-time clinical or educational encounters led or observed by students [32-35, 39-40], reflecting Level 2 of Towle et al’s taxonomy, although in two of these studies the authenticity of the patients is unclear [39–40]. No study reported patient and/or carer involvement above Level 2.

### 4.3 Quality assessment

The MMAT score distribution for the included studies were summarised as follows: low quality n=3 papers [29, 34, 37], moderate quality n=2 [22, 41] and high quality n=15 papers [23-28, 30-32, 33, 35, 36, 38, 39, 40]. The two mixed-methods studies scored 5/5 and 3/5 respectively in the quality criteria for their qualitative and quantitative components respectively [28, 33].

### 4.4 Synthesis of results

A summary of the main results is reported in Table 2. The results have been synthesised in relation to the impact of technology-supported educational activities involving patients and/or carers, on medical students, educators, and patients/ carers themselves. The results are reported in the context of whether studies were low, moderate, or high quality.

**Table 2.**
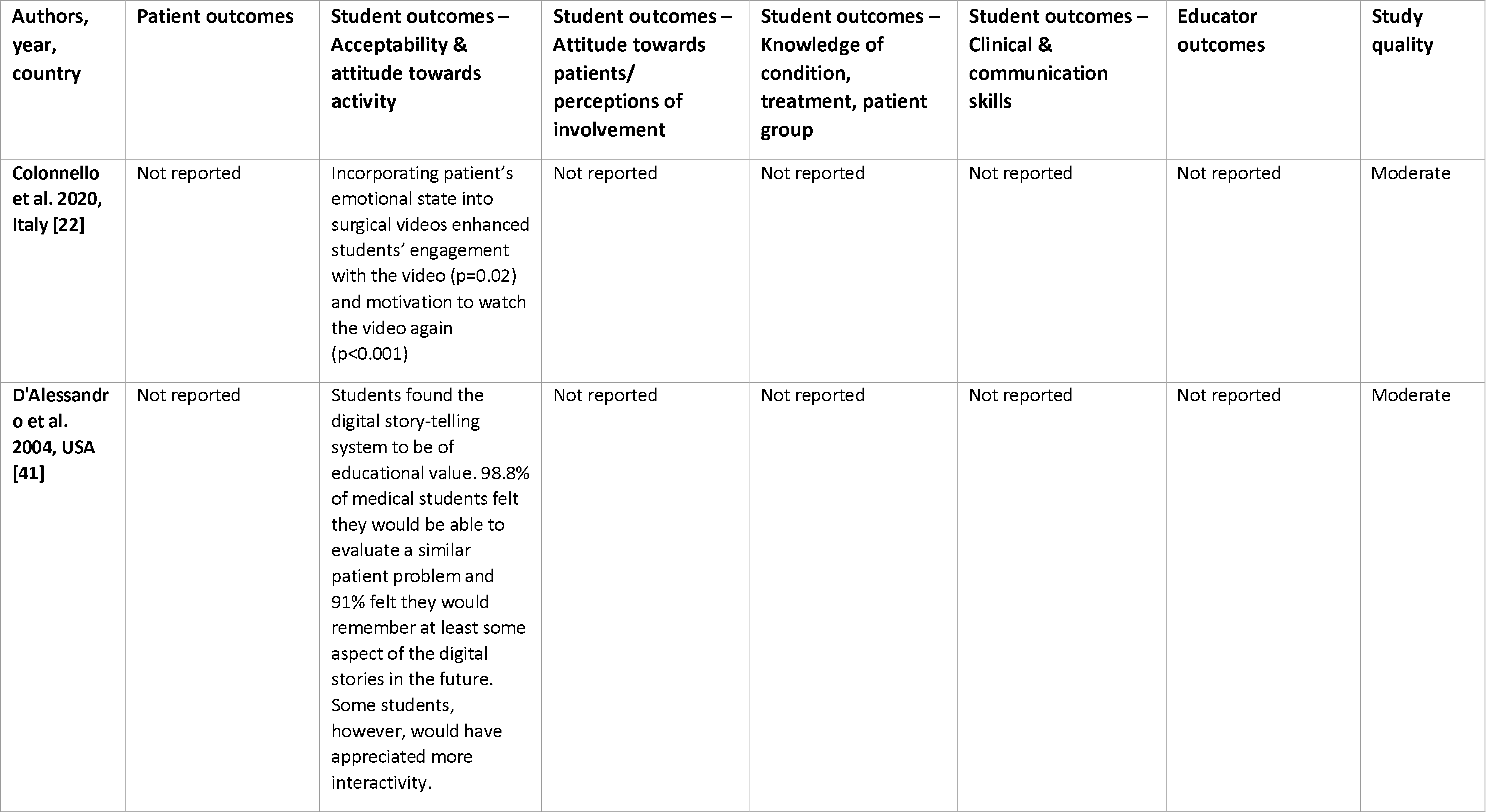
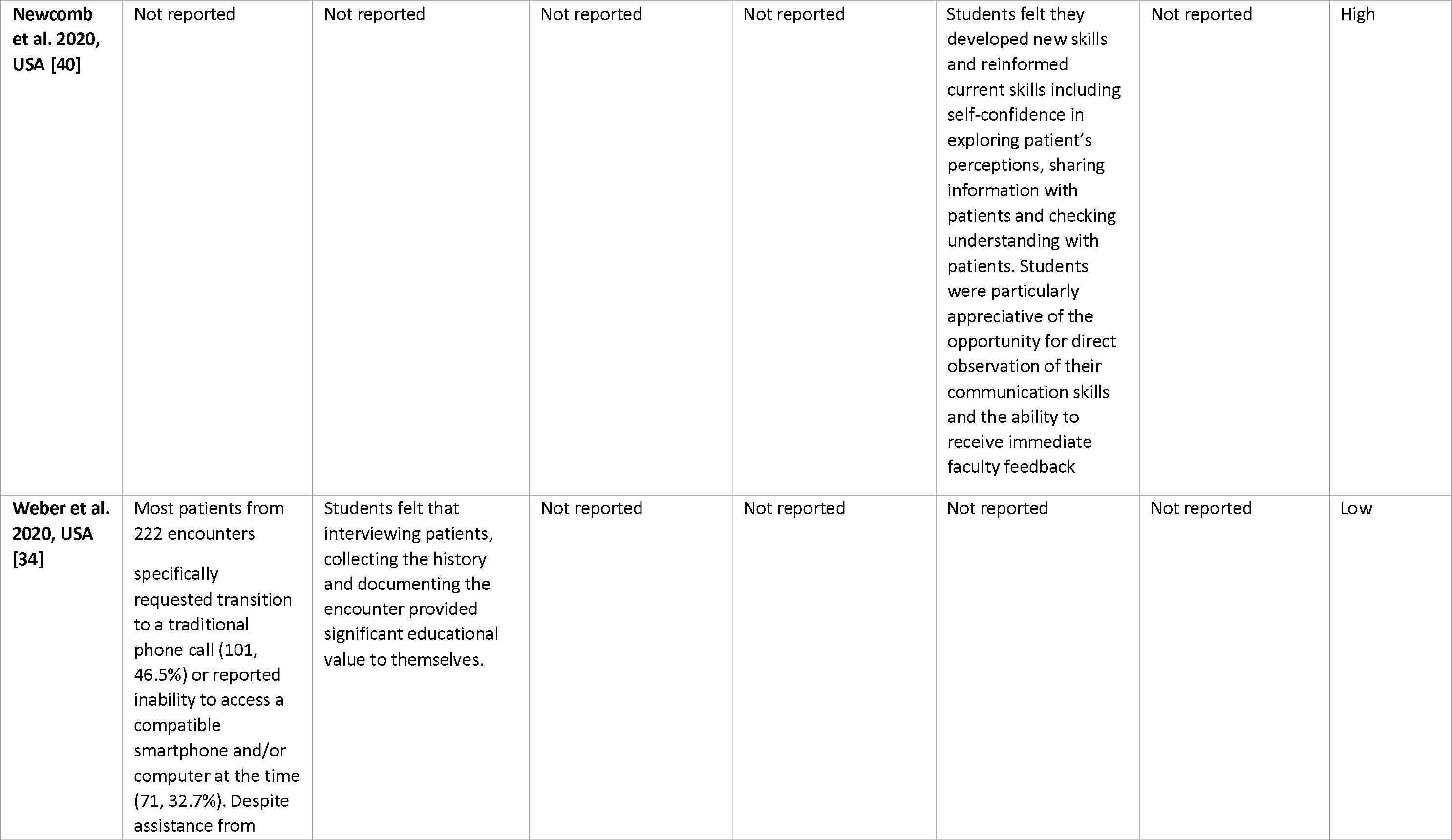
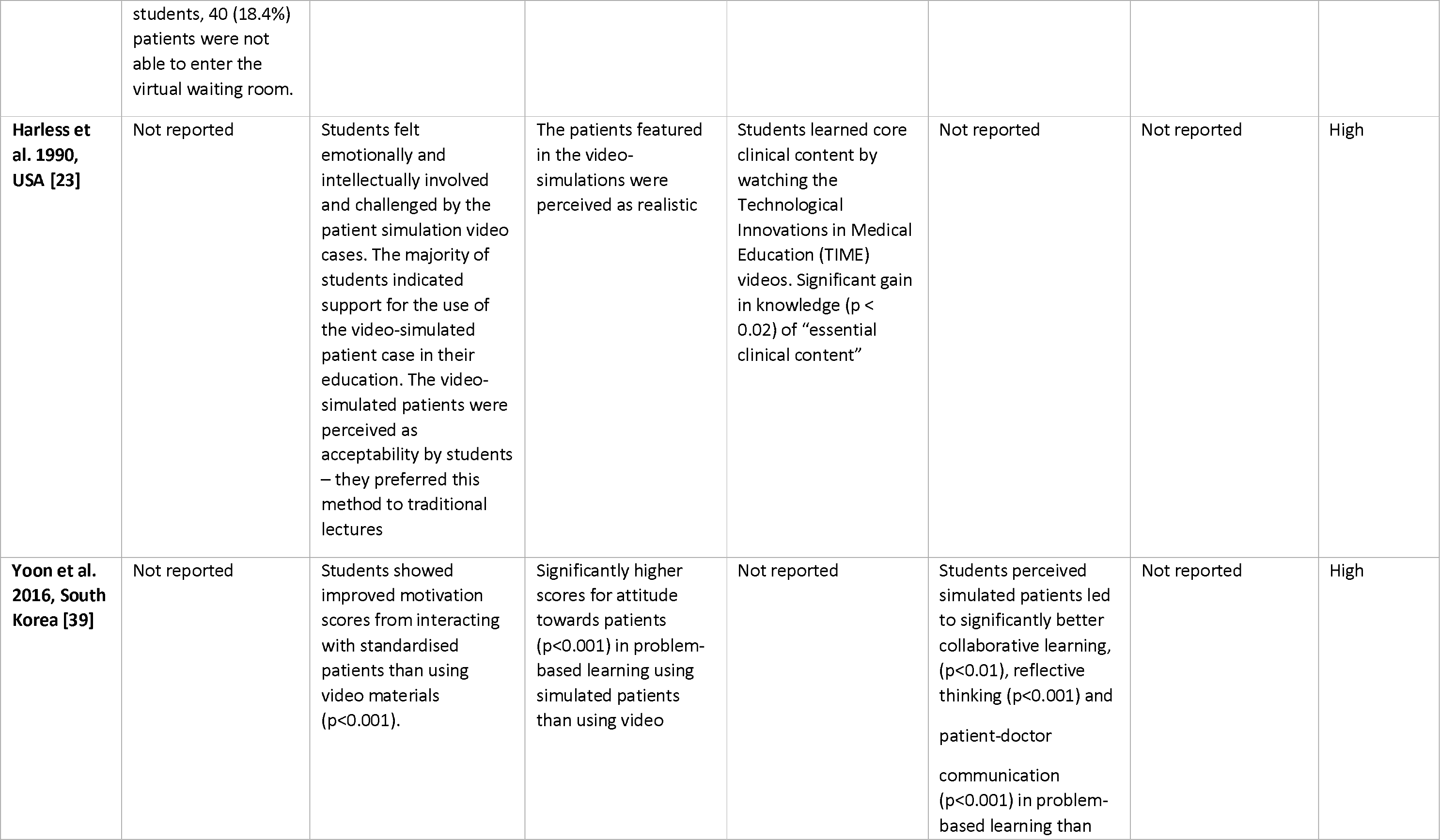
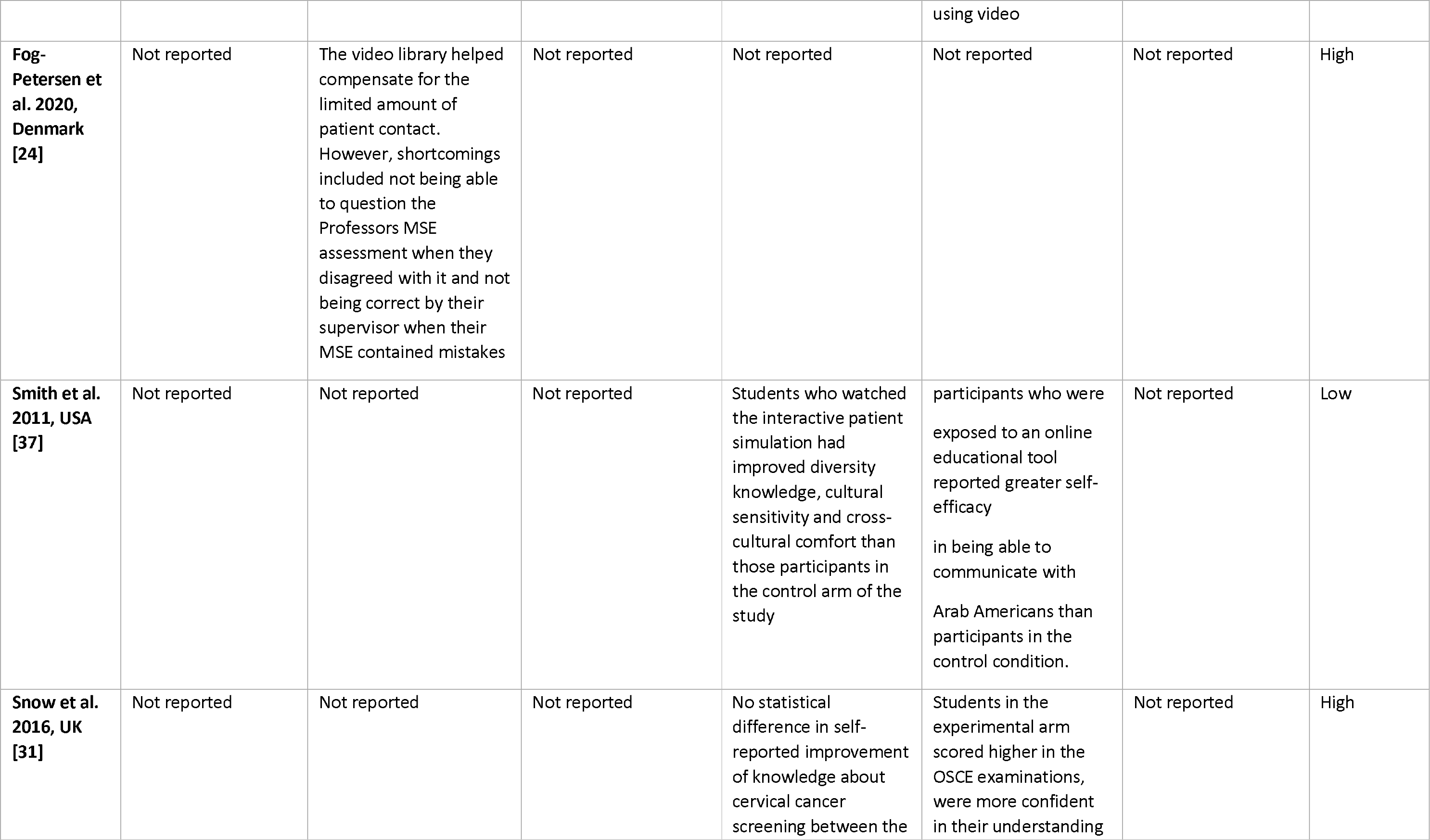
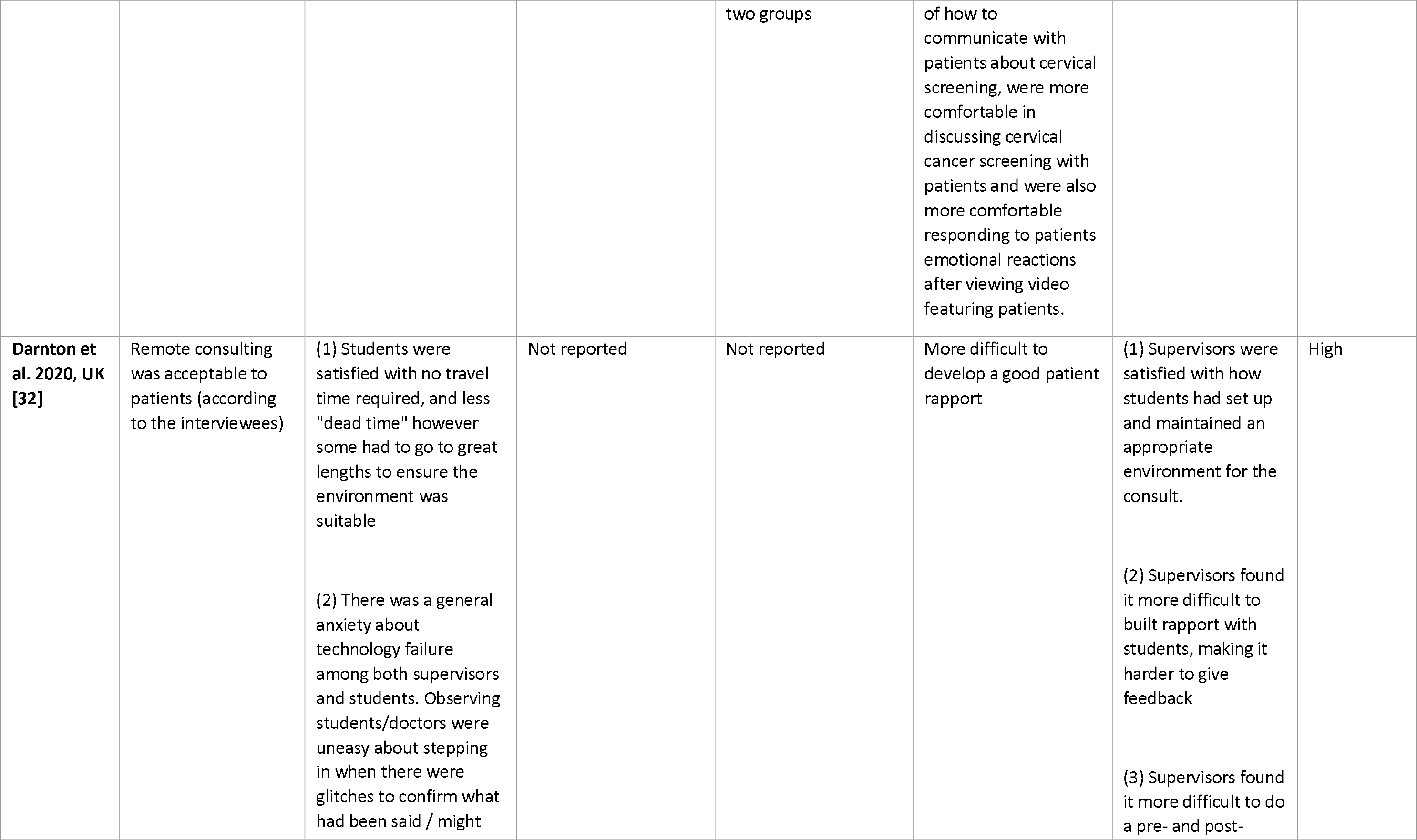
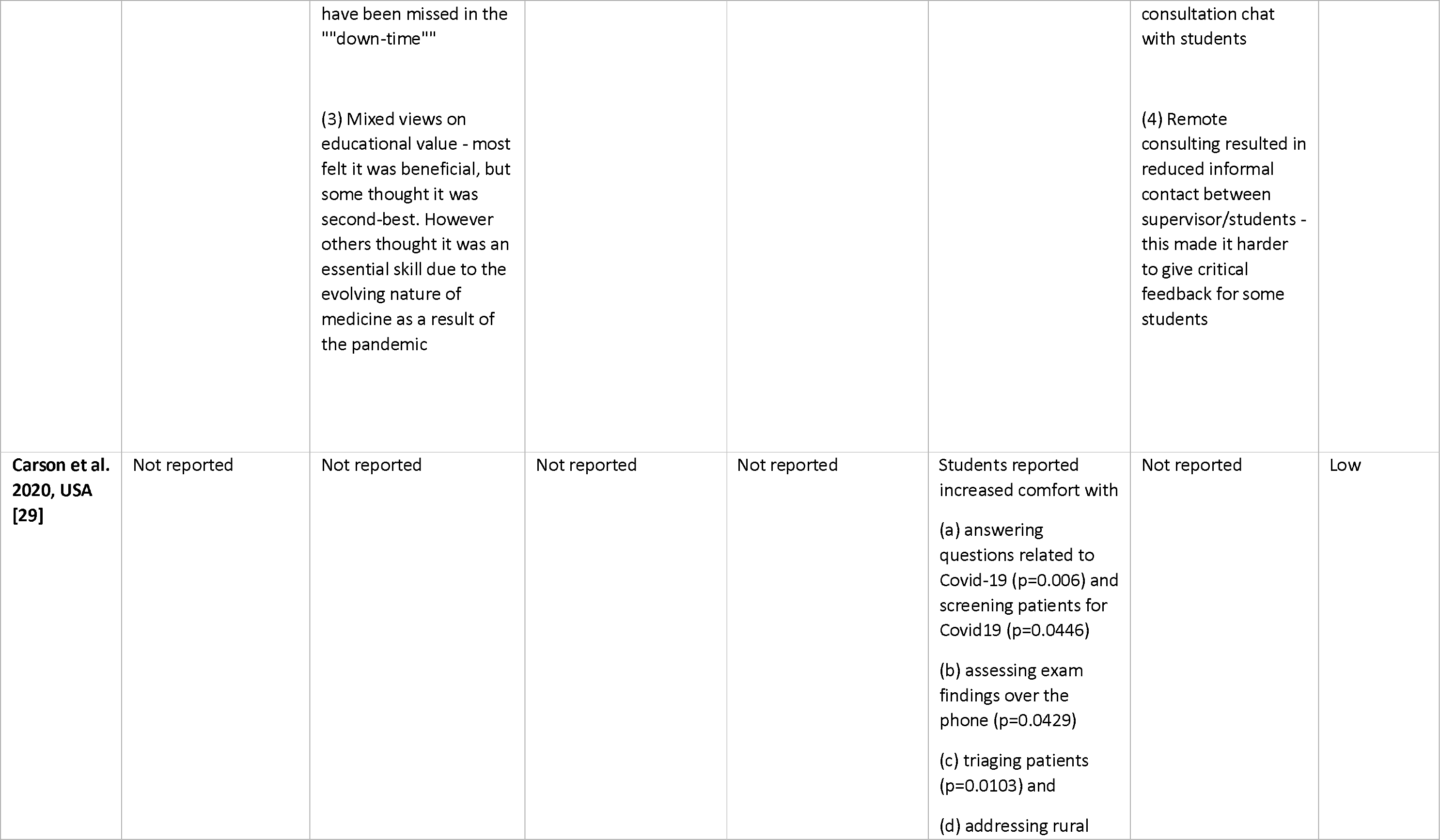
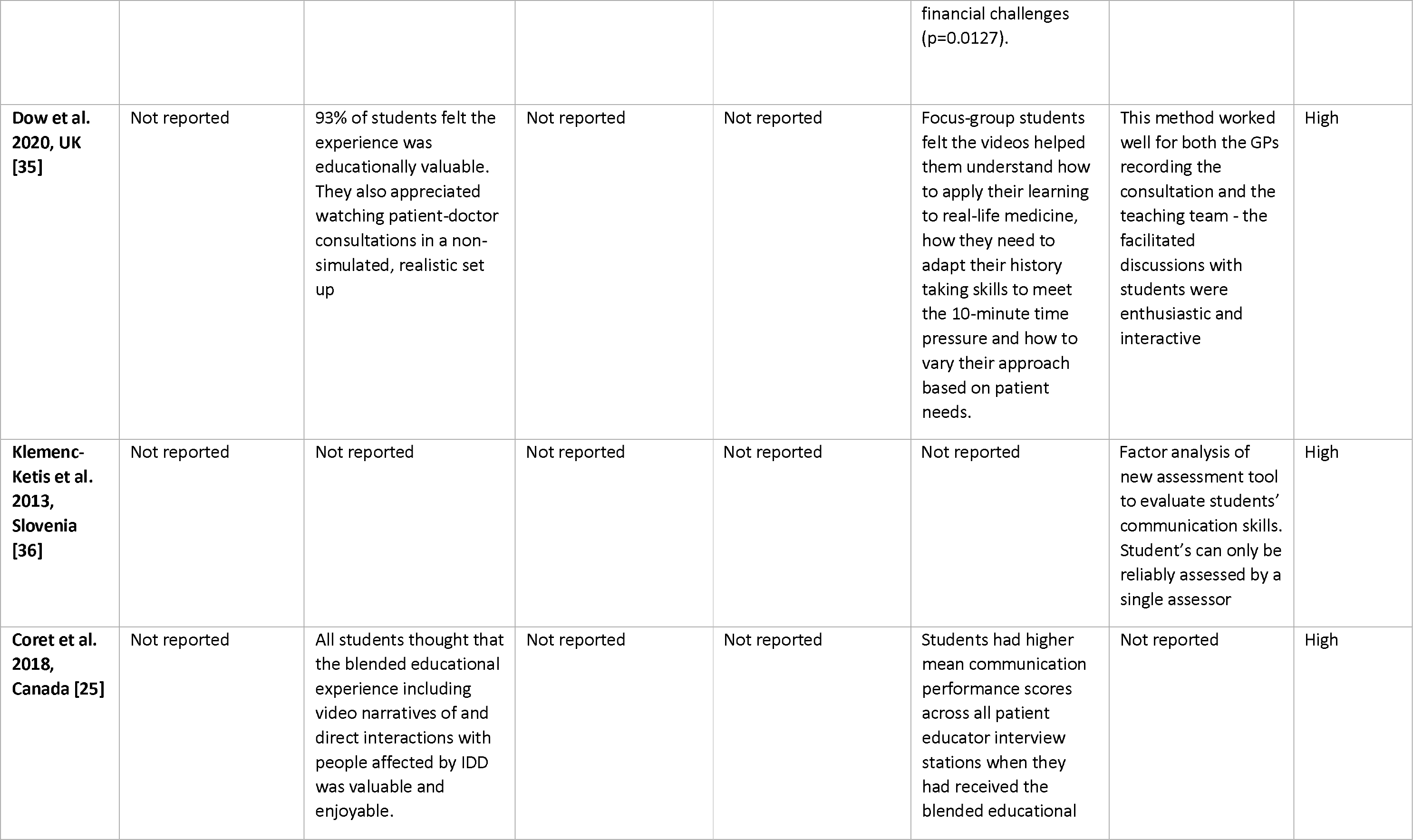
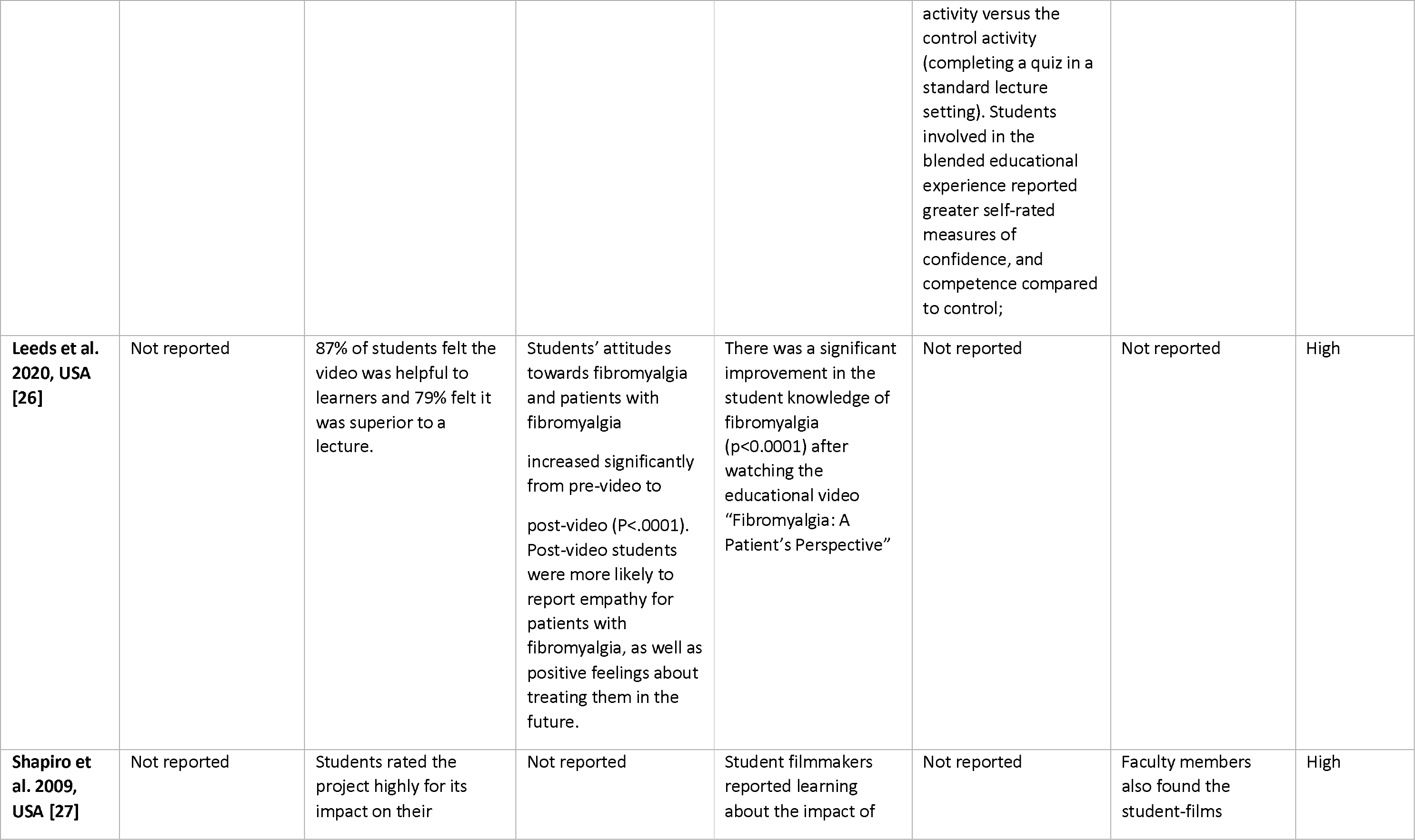
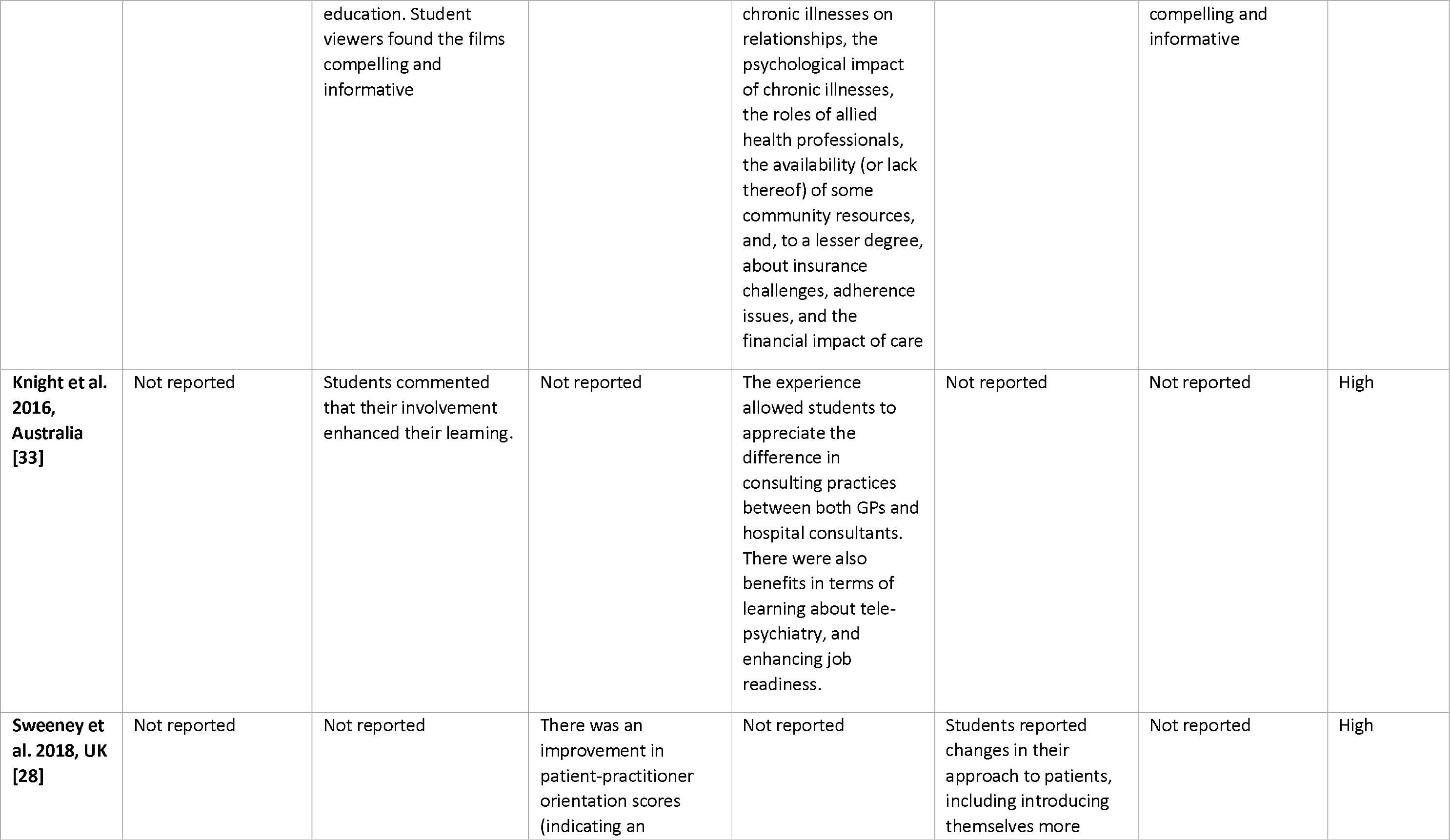
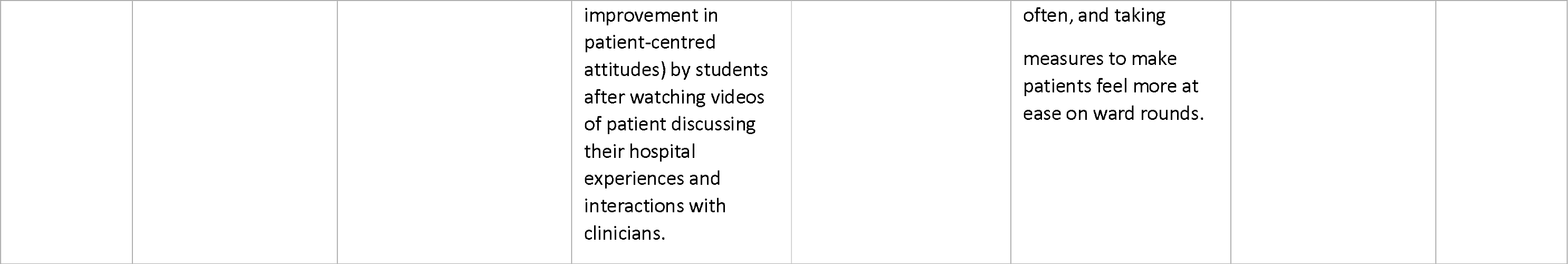
Results of the included studies

#### 4.4.1 Impact on medical students’ learning and attitudes

Nineteen of the 20 articles reported the impact of the technology-supported educational activity on students’ learning and attitudes. Two of these studies included mixed samples of medical students, residents [30, 38] as well as nursing and pharmacy students [30]. We could not differentiate the outcomes for medical students from other healthcare students and so their findings have not been reported in the results described below. The remaining 17 studies measured a variety of medical student-related outcomes, including acceptability and general attitudes towards the educational activity, attitudes towards patients and/or carers, knowledge (of a condition, treatment, or patient group), and clinical and communication skills.

##### 4.4.1.1 Acceptability and general attitudes towards educational activity

Six studies of low to high quality described students reporting technology-supported activities involving patients and/or carers to be educationally valuable [23, 25, 27, 34, 35, 41]. Including an internet-delivered digital story-telling system based on stories and photos shared by patients and families with common paediatric issues [41], a virtual outpatient telehealth clinic delivered by students during the COVID-19 pandemic [34], video-recorded GP consultations [35], videos of patients and caregivers sharing their stories [23, 25, 27]. Two high quality studies reported students found the educational activity acceptable [23, 33]. One high quality study investigating student-led remote consultations found the educational value and acceptability to students was mixed, with some reporting a preference for face to face consultations (e.g. due to being unable to perform a physical examination), while others found the experience valuable [32].

In one high quality study, students reported positive attitudes towards video libraries featuring authentic patient cases [24]. Another high quality study found 79% of students reported that a 13 minute video of a patient’s perspective of fibromyalgia was superior to a traditional face to face lecture [26].

##### 4.4.1.2 Attitudes towards involved patients

Four out of the 20 articles reported students’ attitudes towards patients after the educational activity involving patients or carers. Harless and colleagues [23] found patients featuring in video simulations (where actors had experiences of the issues presented) were perceived by students to be realistic. Two high quality studies found improvement in students’ patient-centred attitudes after watching videos of patients discussing their condition or hospital experiences [26, 28] Yoon and colleagues, however, in a high quality study reported traditional “simulated” patients for problem-based learning led to significantly improved attitude towards patients, compared to a video [39]. However, the reporting of the methods by Yoon and colleagues were limited so it is unclear if patients were authentic in either the standardised or video-delivered approach.

##### 4.4.1.3 Knowledge of condition or treatment featuring in educational activity

Out of six articles reporting students’ knowledge about the condition, treatment, or patient group featuring in technology-supported activities, five reported gains in students’ knowledge. A high quality study reported significant gains in clinical knowledge after viewing a video of simulated patients (based on real patient cases) [23]. A low quality study reported improved knowledge of patient diversity and cultural sensitivity after viewing an online, interactive video of an Arab American Muslim woman receiving care by a white-male GP [37]. Knight and colleagues, in a high quality study, reported the use of telehealth consultations as an educational tool enhanced students’ learning about telepsychiatry as well as their understanding of consulting practices [33]. Two high quality studies reported improved students’ knowledge of patients’ conditions [26–27]. One high quality study reported no differences in self-reported knowledge about cervical screening when students viewed a video involving patients, versus a video featuring a clinician [31].

##### 4.4.1.4 Clinical and communication skills

Six studies reported improvements in students’ communication skills after participating in technology-supported teaching involving patients and/or carers. One high quality qualitative study reported a class delivered via video-conferencing with “simulated” patients helped students develop skills in exploring patient’s perceptions, sharing information with patients, and checking understanding [40]. Participants exposed to an online educational tool featuring a Muslim woman reported greater self-efficacy in being able to communicate with Arab American patients than participants in the control condition [37]. In their high quality qualitative study, Dow and colleagues [35] reported videos helped medical students understand how to adapt their history taking skills and vary their approach based on patient’s needs. Coret and colleagues [25], in a high quality study, reported higher communication scores when students received a blended educational activity versus a standard lecture setting. Students reported introducing themselves more often, and taking measures to make patients feel more at ease, after watching videos of patients discussing their hospital experiences in a high quality study by Sweeney et al [28]. Snow et al [31] reported students felt more confident communicating with patients about cervical screening, and more comfortable responding to patients’ emotional reactions, after watching a video of patients sharing their experiences of colposcopy, compared to their peers who watched a video featuring a clinician only.

One low quality study found a student-led hotline for patients with COVID-19 increased students’ remote clinical skills in screening, assessment, and triaging patients [29]. Snow and colleagues [31] reported higher student OSCE scores after watching the patient video, compared with students who watched the clinician-only video.

However, the traditional standardised patient simulation employed by Yoon and colleagues [39] was found to be significantly more beneficial to students in their collaborative learning, reflective thinking, and patient-doctor communication, than the video-delivered simulation. Further, student-led remote consultations were reported by some students to inhibit rapport-building with patients versus traditional face to face consultations [32].

#### 4.4.2 Perspective of medical educators

##### 4.4.2.1 Acceptability and value of educational activity

In Dow and colleagues’ study of video-recorded GP consultations, medical teachers reported the recordings facilitated discussions with students and provided an interactive teaching session [35]. Shapiro et al [27] found films made by students about the impact of living with chronic conditions to be compelling and informative from the perspective of their tutors. While GP supervisors observing student-led remote patient consultations were satisfied with how students set up and maintained appropriate environments for consultations, the physical distance between them and the student made it more difficult to build rapport with students, with fewer opportunities to offer students feedback [32].

##### 4.4.2.2 Perceptions of students’ skills

One high quality study developed a new tool for educators to assess students’ communication skills on medical questions asked by patients in a virtual setting (internet e-forum) [36], finding the e-forum questions from patients to be a suitable learning tool for assessing clinical decision-making in medical students [36].

#### 4.4.3 Perspective of patients and/ or carers

##### 4.4.3.1 Acceptability of educational activity

Darnton and colleagues [32] reported that student-led remote consultations were acceptable to patients, although this was anecdotal evidence from the perspective of students and educators.

##### 4.4.3.2 Barriers to participating in educational activity

Weber and colleagues [34] reported some difficulties for patients in attempting to participate in student-led telehealth consultations, via the students who took part. Out of 222 encounters, 46.5% of patients requested a traditional telephone call (over the telehealth consultation), 32.7% reported not having access to a compatible smartphone and/or computer and 18.4% had difficulty with the technology and were unable to join the virtual waiting room [34].

## 5. DISCUSSION

### 5.1 Main findings

The aim of this rapid systematic review was to identify evidence for the use of technology in undergraduate medical education when patients and/or carers were involved, the levels of involvement in this context, and what barriers and facilitators to their involvement have been reported. Twenty articles met the eligibility criteria and demonstrated a variety of undergraduate medical education settings in which technology can be used, without excluding the involvement of patients and caregivers. However, levels of involvement were generally low and no study reported involvement above Level 2.

The review found that technology-supported teaching sessions involving patients and/or carers were found to be educationally valuable to students and educators, were acceptable to students, and increased students’ knowledge of patient groups, as well as their communication and clinical skills. Limited evidence also demonstrated enhanced student engagement, and improved patient-centred attitudes. Two studies highlighted the challenges associated with technology-delivered medical education when students were involved in remote clinical encounters including difficulty building rapport with patients, and between GP supervisors and students [32], and the potential for patients to lack access to suitable technology to be able to engage in telehealth when students are involved [34].

### 5.2 Links to previous research

No study directly captured the views of patients or carers participating in technology-delivered medical education. This lack of insight from patients and carers is unusual in the context of a growing body of literature reporting PPI in medical education generally [3]. Such research has highlighted ways patients and carers wish to be involved, including wanting clear information before any student encounter and a desire for their consent to be taken at each stage (e.g. may consent to student being present, but not taking a clinical examination) [42, 43]. The use of technology as an educational tool has significantly increased since the beginning of the COVID-19 pandemic [8, 11, 44]. Interactive technologies (e.g. video-conferencing software) provide students with valuable experiences interacting with patients and carers in real-time [11, 44]. It would therefore be beneficial to understand patients’ opinions about taking part in remote interactions with students, to understand further how their involvement can be supported, fully consented, and what challenges or opportunities may arise when technology is used. One study in our review highlighted the potential for patients and carers to be digitally excluded from medical education when sessions involve technology [34], which is in line with recent research [13].

There was poor consistency in the use of terminology to describe involved patients and carers. Studies using the terms “virtual patient”, “simulated patient”, or “standardized patient” tended to assume meaning was understood and no description was offered. Previous authors have highlighted the inconsistencies in meaning within and between these common terms [45, 46]. This has implications for the replicability of medical education research evaluating the involvement of patients and/or carers. Going forward, we recommend that researchers and educationalists standardise the use of terms using available definitions. For instance, Towle and colleagues clearly differentiate “patients” (who have a medical problem), from “simulated/ standardised” patients who role play symptoms and signs they do not actually have [5]. We would argue as these are not patients, the term “patients” should be avoided, instead using more descriptive terms e.g. “actors”.

Finding low levels of involvement in the studies reviewed is also inconsistent with literature evaluating PPI in undergraduate medical education within in-person settings. The majority of studies in a recent systematic review described patients as educators and assessors, reflecting Level 4 of Towle’s taxonomy [3, 5]. Some studies in our review involved patients with real experience of the medical problems they portrayed, but scripted their role, supposedly with the aim of standardising students’ learning experience [23]. A potential benefit of doing this might be to provide students with a “safe” environment in which to make mistakes [47]. Whilst this may be a valid approach to use in early years of student training, scripted, “virtual” or “simulated” patients can be perceived as formulaic by medical students, failing to prepare them well for dealing with issues beyond the clinical presentation, such as unemployment, disability, or other life circumstances [48]. Remote teaching opportunities with authentic patients would ensure medical students gain remote consultation skills, and prepare them for the wider issues that real patient encounters bring. Future research should capture the demographic of patients or carers who are willing and able to use the technology to ensure patient encounters are representative of the populations medical students serve.

### 5.4 Limitations

This was a rapid systematic review, conducted under time constraints and we acknowledge the potential to have excluded some relevant research, e.g. foreign language articles. We acknowledge the inclusion of four studies where it remains unclear whether authentic patients or carers were involved, due to poor describing of methods. We decided to retain these studies as there was also no indication that patients and/or carers were not authentic. This raises an important issue within the literature whereby a lack of description inhibits a thorough assessment and, indeed, replication, of the study methods. We acknowledge that our PPI contributors were not involved in refining the research question, however their contribution to the review processes, and to our understanding of issues related to whether “authentic” patients were involved in educational activities or not, as described by study authors, was invaluable.

## 6. CONCLUSIONS

Medical schools should ensure students’ learning is reflective of everyday healthcare practice during the COVID-19 pandemic and beyond, by incorporating patient and carer involvement in remote teaching sessions using a broad array of technologies. Technology-supported teaching sessions involving patients and/or carers were found to be acceptable to students, improved their patient-centred attitudes and skills, as well as their knowledge of patient groups. Limited evidence highlighted challenges associated with remote medical education when students were involved in clinical encounters, including difficulty building rapport between students and patients, and GP supervisors and students, and the potential for patients to lack access to suitable technology to be able to engage in the interaction. With the majority of studies describing patient and carer involvement at low levels, there is a need for medical education teams to evaluate higher levels of patient and carer involvement (e.g. design, delivery and evaluation of medical curricula), and to capture patients’ perceptions of being involved in teaching (including barriers and facilitators) when using technologies related to recent advances in healthcare delivery (e.g. telehealth). Overall, quality of the studies included in this review was moderate to high; the results of studies of poor quality and those lacking clear descriptions of patients and carers should be viewed with caution.

## Data Availability

Not applicable

## Acknowledgments

This review was funded by the National Institute for Health Research (NIHR) School for Primary Care (SPCR) and University College London (UCL). We’d also like to acknowledge the support of Claire Duddy at Nuffield Department of Primary Care Health Sciences, University of Oxford, for assistance with sourcing full text articles.

## Supplementary Materials

The following are available online at [link to be confirmed]

## Author Contributions

Study design: SLW, AA, JHH, NY, CJ, NC, SP; Searches: NR, SLW; Data acquisition: SLW, EL, AA, JHH, NY, NR, AA, CJ, NC; Data analysis: SLW, EL, AA, JHH, NY, CJ, NC, SP; Initial draft manuscript: SLW, EL; Reviewing and editing manuscript: SLW, AA, JHH, NY, CJ, NC, NR, SP. All authors have read and agreed to the published version of the manuscript.

## Ethical Approval

Not applicable

## REFERENCES

1. National Health Service, The NHS long term plan. 2019, NHS England London.

2. Council, G.M., Patient and public involvement in undergraduate medical education, in Tomorrow’s Doctors (2009). 2011, General Medical Council.

3. Dijk, S.W., E.J. Duijzer, and M. Wienold, Role of active patient involvement in undergraduate medical education: a systematic review. BMJ open, 2020. 10(7): p. e037217.

4. Stacy, R. and J. Spencer, Patients as teachers: a qualitative study of patients’ views on their role in a community-based undergraduate project. Medical education, 1999. 33(9): p. 688–694.

5. Towle, A., et al., Active patient involvement in the education of health professionals. Medical education, 2010. 44(1): p. 64–74.

6. Spencer, J., et al., Patient-oriented learning: a review of the role of the patient in the education of medical students. Medical education, 2000. 34(10): p. 851–857.

7. Tew, J., C. Gell, and S. Foster, Learning from Experience: Involving service users and carers in mental health education. 2004.

8. Paul, N., et al., Integration of Technology in Medical Education on Primary Care During the COVID-19 Pandemic: Students’ Viewpoint. JMIR Medical Education, 2020. 6(2): p. e22926.

9. Richards, T. and H. Scowcroft, Patient and public involvement in covid-19 policy making. 2020, BMJ.

10. Greenhalgh, T., et al., Video consultations for covid-19: An opportunity in crisis? 2020, BMJ.

11. Monaghesh, E. and A. Hajizadeh, The role of telehealth during COVID-19 outbreak: a systematic review based on current evidence. BMC Public Health, 2020. 20(1): p. 1–9.

12. Munday, D., et al., Using Zoom: Views from Patient and Public Involvement Contributors. 2020, NIHR Applied Research Collaboration East of England.

13. Greer, B., et al., Digital exclusion among mental health service users: qualitative investigation. Journal of medical Internet research, 2019. 21(1): p. e11696

14. Gordon, M., et al., Patient/service user involvement in medical education: A best evidence medical education (BEME) systematic review: BEME Guide No. 58. Medical Teacher, 2020. 42(1): p. 4–16.

15. Dogba, M.J., et al., Using information and communication technologies to involve patients and the public in health education in rural and remote areas: a scoping review. BMC health services research, 2019. 19(1): p. 128.

16. Varker, T., et al., Rapid evidence assessment: increasing the transparency of an emerging methodology. Journal of evaluation in clinical practice, 2015. 21(6): p. 1199–1204.

17. Ouzzani, M., et al., Rayyan — a web and mobile app for systematic reviews. Systematic Reviews, 2016. 5.

18. Cooke, A., D. Smith, and A. Booth, Beyond PICO: the SPIDER tool for qualitative evidence synthesis. Qualitative health research, 2012. 22(10): p. 1435–1443

19. Hong, Q.N., et al., Mixed methods appraisal tool (MMAT), version 2018. Registration of copyright, 2018. 1148552.

20. Ghio, D., et al., What Influences People’s Responses To Public Health Messages For Managing Risks and Preventing Disease During Public Health Crises? A Rapid Review of the Evidence and Recommendations. PsyArXiv Preprints, 2020.

21. Lawes-Wickwar, S., et al., A rapid systematic review of public responses to health messages encouraging vaccination against infectious diseases in a pandemic or epidemic. Vaccines, 2021. 9(2): p. 72.

22. Colonnello, V., et al., Emotionally salient patient information enhances the educational value of surgical videos. Advances in Health Sciences Education, 2020. 25(4): p. 799–808.

23. Harless, W.G., et al., A field test of the TIME patient simulation model. Academic Medicine, 1990. 65(5): p. 327–33.

24. Fog-Petersen, C., et al., Clerkship students’ use of a video library for training the mental status examination. Nordic Journal of Psychiatry, 2020. 74(5): p. 332–339.

25. Coret, A., et al., Patient Narratives as a Teaching Tool: A Pilot Study of First-Year Medical Students and Patient Educators Affected by Intellectual/Developmental Disabilities. Teaching & Learning in Medicine, 2018. 30(3): p. 317–327.

26. Leeds, F.S., et al., A Patient-Narrative Video Approach to Teaching Fibromyalgia. Journal of Medical Education & Curricular Development, 2020. 7: p. 2382120520947068.

27. Shapiro, D., L. Tomasa, and N.A. Koff, Patients as teachers, medical students as filmmakers: the video slam, a pilot study. Academic Medicine, 2009. 84(9): p. 1235–43.

28. Sweeney, K. and P. Baker, Promoting empathy using video-based teaching. The clinical teacher, 2018. 15(4): p. 336–340.

29. Carson, S., et al., Student Hotline Improves Remote Clinical Skills and Access to Rural Care. PRiMER: Peer-Review Reports in Medical Education Research, 2020. 4.

30. Gorniewicz, J., et al., Breaking bad news to patients with cancer: a randomized control trial of a brief communication skills training module incorporating the stories and preferences of actual patients. Patient education and counseling, 2017. 100(4): p. 655–666.

31. Snow, R., et al., Does hearing the patient perspective improve consultation skills in examinations? An exploratory randomized controlled trial in medical undergraduate education. Medical teacher, 2016. 38(12): p. 1229–1235

32. Darnton, R., et al., Medical students consulting from home: A qualitative evaluation of a tool for maintaining student exposure to patients during lockdown. Medical Teacher, 2020: p. 1–8.

33. Knight, P., et al., Positive Clinical Outcomes Are Synergistic With Positive Educational Outcomes When Using Telehealth Consulting in General Practice: A Mixed-Methods Study. Journal of Medical Internet Research, 2016. 18(2): p. e31.

34. Weber, A.M., et al., An outpatient telehealth elective for displaced clinical learners during the COVID-19 pandemic. BMC medical education, 2021. 21(1): p. 1–8.

35. Dow, N., et al., ‘GP Live’-recorded General Practice consultations as a learning tool for junior medical students faced with the COVID-19 pandemic restrictions. Education for Primary Care, 2020. 31(6): p. 377–381.

36. Klemenc-Ketis, Z. and J. Kersnik, New virtual case-based assessment method for decision making in undergraduate students: a scale development and validation. BMC Medical Education, 2013. 13: p. 160.

37. Smith, B.D. and K. Silk, Cultural competence clinic: an online, interactive, simulation for working effectively with Arab American Muslim patients. Academic Psychiatry, 2011. 35(5): p. 312–6.

38. Kindratt, T., et al., Parent-provider paediatric literacy communication: A curriculum for future primary care providers. Perspectives on Medical Education, 2019. 8(2): p. 110–117.

39. Yoon, B.Y., et al., Using standardized patients versus video cases for representing clinical problems in problem-based learning. Korean Journal of Medical Education, 2016. 28(2): p. 169–78.

40. Newcomb, A.B., et al., Building Rapport and Earning the Surgical Patient’s Trust in the Era of Social Distancing: Teaching Patient-Centered Communication During Video Conference Encounters to Medical Students. Journal of Surgical Education., 2020.

41. D’Alessandro, D.M., T.E. Lewis, and M.P. D’Alessandro, A pediatric digital storytelling system for third year medical students: the virtual pediatric patients. BMC Medical Education, 2004. 4: p. 10.

42. Gandsas, A., et al., Live streaming video for medical education: a laboratory model. Journal of Laparoendoscopic & Advanced Surgical Techniques. Part A, 2002. 12(5): p. 377–82.

43. Alao, A., et al., Real-time patients’ perspectives about participating in teaching consultations in primary care: A questionnaire study. Medical Teacher, 2021: p. 1–21.

44. Major, C., Innovations in Teaching and Learning during a Time of Crisis. Innovative Higher Education, 2020. 45: p. 265–266.

45. Kononowicz, A.A., et al., Virtual patients-what are we talking about? A framework to classify the meanings of the term in healthcare education. BMC medical education, 2015. 15(1): p. 1–7.

46. Adamo, G., Simulated and standardized patients in OSCEs: achievements and challenges 1992-2003. Medical Teacher, 2003. 25(3): p. 262–70.

47. Hege, I., et al., How to tell a patient’s story? Influence of the case narrative design on the clinical reasoning process in virtual patients. Medical teacher, 2018. 40(7): p. 736–742.

48. Urresti-Gundlach, M., et al., Do virtual patients prepare medical students for the real world? Development and application of a framework to compare a virtual patient collection with population data. BMC Medical Education, 2017. 17(1): p. 174.

