## Supplementary File 1 for "Technology-delivered undergraduate medical education involving patients and carers: A rapid systematic review"

Supplementary File 1: Search Strategy (Ovid MEDLINE)

|  | **Searches** |
| --- | --- |
| 1 | Students, Medical/ |
| 2 | education, medical/ or education, medical, undergraduate/ |
| 3 | ((medical or medicine) adj2 (student? or undergraduate?)).ti,ab,kw. |
| 4 | medical education.ti. |
| 5 | 1 or 2 or 3 or 4 |
| 6 | communications media/ or blogging/ or social media/ or exp telecommunications/ or exp telephone/ or exp videoconferencing/ or wireless technology/ |
| 7 | (digital* or electronic* or virtual* or web* or internet* or online* or video* or technolog*).ti. |
| 8 | ((digital* or electronic* or virtual* or web* or internet* or online* or video* or remote or tele*) adj3 (meeting? or attend* or conferenc* or class* or workshop* or seminar* or tutorial* or module*)).ti,ab,kw. |
| 9 | (((online or electronic) adj learning) or (elearning or e-learning)).ti,ab,kw. |
| 10 | (youtube* or vlog* or blog* or social media or discussion group? or facebook or email* or e-mail* or textmessag* or text-messag* or sms or twitter or tweet* or instagram or whatsapp or whats app or skype or zoom or MS Teams or microsoft teams or teleconf* or live stream* or livestream*).ti,ab,kw. |
| 11 | ((digital* or electronic* or virtual* or web* or internet* or online* or video* or remote or tele* or technolog*) adj (enabled or driven or based)).ti,ab,kw. |
| 12 | 6 or 7 or 8 or 9 or 10 or 11 |
| 13 | (adult children/ or caregivers/ or exp disabled persons/ or grandparents/ or exp legal guardians/ or exp parents/ or patients/ or siblings/ or spouses/ or exp survivors/) and (engag* or participat* or collaborat* or involv* or codesign* or co-design* or coproduc* or co-produc*).ti,ab,kw. |
| 14 | Patient Participation/ |
| 15 | Community Participation/ |
| 16 | ((patient? or client? or consumer? or user? or survivor?) adj5 (engag* or participat* or collaborat* or involv* or codesign* or co-design* or coproduc* or co-produc*)).ti,ab,kw. |
| 17 | ((carer? or care giver? or caregiver? or family or parent or parents or guardian? or spouse? or husband? or wives or father? or mother? or parent? or brother? or sister? or sibling?) adj5 (engag* or participat* or collaborat* or involv* or codesign* or co-design* or coproduc* or co-produc*)).ti,ab,kw. |
| 18 | ((public or general population) adj5 (engag* or participat* or collaborat* or involv* or codesign* or co-design* or coproduc* or co-produc*)).ti,ab,kw. |
| 19 | (patient? adj2 (teacher? or trainer? or educator? or leader?)).ti,ab,kw. |
| 20 | ((patient centered or patient centred or patient focused or patient driven or patient initiated) adj educat*).ti,ab,kw. |
| 21 | (patient adj2 (narative? or stories or storytell* or autobiograph* or biograph*)).ti,ab,kw. |
| 22 | 13 or 14 or 15 or 16 or 17 or 18 or 19 or 20 or 21 |
| 23 | 5 and 12 and 22 |
